# BREASTFEEDING SUPPORT BASED ON SELF-CARE FORM PERSPECTIVE PHILOSOPHY OF SCIENCE: A LITERATURE REVIEW

**DOI:** 10.1101/2024.11.16.24315829

**Authors:** Sumirah Budi Pertami, Moses Glorino Rumambo Pandin, Esti Yunitasari, Mira Triharini

## Abstract

**Introduction:** Failure of the lactation process leads to decreased milk production, which is a serious problem especially regarding the health of the mother and child. Reduced milk production and failure to provide exclusive breastfeeding directly contribute to high infant morbidity.. Infants who are not breastfed can deteriorate their health and increase the risk of malnutrition and stunting. Breastfeeding mothers need the support of those closest to them, such as family members, friends, relatives and coworkers. The family, in this case the husband or parents, is considered the most capable party to influence the mother to maximize exclusive breastfeeding. Support from others or those closest to them plays a significant role in the success or failure of breastfeeding. The purpose of this study is to examine the philosophy of breastfeeding support from the perspective of the Self Care model theory.

**Methods:** This study used a literature review using the PICOS framework. Articles were searched from 3 databases namely Sciencedirect, Scopus, Sage. The keywords used in the literature search were breastfeeding AND support, AND self AND care AND theory AND model, AND postpartum. The search was limited to 2020-2024 publications, full articles, not review articles. The strategy used to search for articles using the PICOS framework. Then selected using the PRISMA diagram and obtained eleven articles. Quality assessment of eleven articles using the JBI Critical Appraisal tool.

**Results:** Eleven articles describing the theoretical application of the Dhorothy Orem Self Care Model can be used as guidelines in providing self-care to improve breastfeeding support.

**Conclusion:** a more efficient strategy to increase breast milk production can be developed by understanding the self-care model. In addition, the self-care model can be applied in breasfeeding support by providing self-care to improve breastfeeding practices.

## INTRODUCTION

Babies get the best nutrition from breast milk. Exclusive breastfeeding is breast milk given exclusively for six months without additional food or fluids. (Thet et al., 2016). Early initiation of breastfeeding (IMD), exclusive breastfeeding for the first six months of life and continued breastfeeding until 24 months of age are important ways to reduce infant mortality, especially in developing countries according to the World Health Organization (WHO) and United Nations Children’s Fund (UNICEF) (Kumar, 2014). Exclusive breastfeeding has many health and economic benefits for mothers, infants, and society as a whole (Freney et al., 2016). There is some evidence that exclusive breastfeeding reduces health costs, affecting both developed and developing economies. However, this limited evidence does not suggest that policymakers in developed and developing countries should prioritize breastfeeding (Freney et al., 2016). Breastfeeding rates are increasing very slowly. However, according to (Gupta, 2019) pressure on the supply of breastmilk substitutes will increase worldwide.

Exclusive breastfeeding rates are low worldwide. Only 43% of newborns receive early initiation of breastfeeding (IMD), and 40% are exclusively breastfed (Kumar, 2014). The percentage of breastfeeding starts less than one hour in Indonesia is still very low at 34.5%. West Papua Province had the lowest percentage at 21.7%, followed by Riau Province at 22.1%, and Riau Islands at 22.7%.(Kemenkes, 2013). Delaying breastfeeding will increase morbidity and mortality. In Central Java in 2017, 63.93% of under-fives (children under 2 years old) were exclusively breastfed. Most of the urban infants were exclusively breastfed, while 65.56% were exclusively breastfed in rural areas (Kemenkes, 2013).

Ideal breast milk production is very important for the health of both mother and baby, as breast milk contains essential nutrients and immune substances that protect babies from various diseases. Although breastmilk is widely recognized as a nutrient, some mothers are unable to produce enough milk. Family is often critical to successful breastfeeding. More efficient strategies to increase breast milk production can be developed by understanding family support through the self-care model, which focuses on empowering individuals to take control of their own health; in this context, systematic and integrated family support is required. Although they have been informed about the benefits of breastmilk and appropriate breastfeeding methods, many mothers have difficulty producing enough milk. This is a common phenomenon. Factors such as stress, lack of knowledge about effective breastfeeding methods, and inadequate social support are some that often affect this difficulty. It can be even worse if you don’t have enough family support. This fact suggests that there needs to be a more holistic approach to helping breastfeeding mothers, where the family actively and effectively participates in assisting the breastfeeding process.. According to (Roesli, 2007), families provide the most support for the success of exclusive breastfeeding. Breastfeeding mothers need support and assistance, both when starting breastfeeding and after two years. This support is especially needed from family members, especially husbands, and medical personnel (Proverawati, 2010).

Support from others and loved ones is essential for successful breastfeeding. The ability to maintain breastfeeding will increase if there is support. A mother who does not get support from her husband, mother, sister, or even fear is influenced to switch to formula milk (Proverawati, 2010).

## MATERIALS AND METHODS

The literature for the originality of this study used articles in English from 3 databases: Sciencedirect, Scopus, Sage. The keywords used in the literature search were: breastfeeding AND support, AND self AND care AND theory AND model, AND postpartum. The search was limited to 2020-2024 publications, full articles, not review articles. The strategy used to search for articles using the PICOS framework consists of:

**Figure 1.**
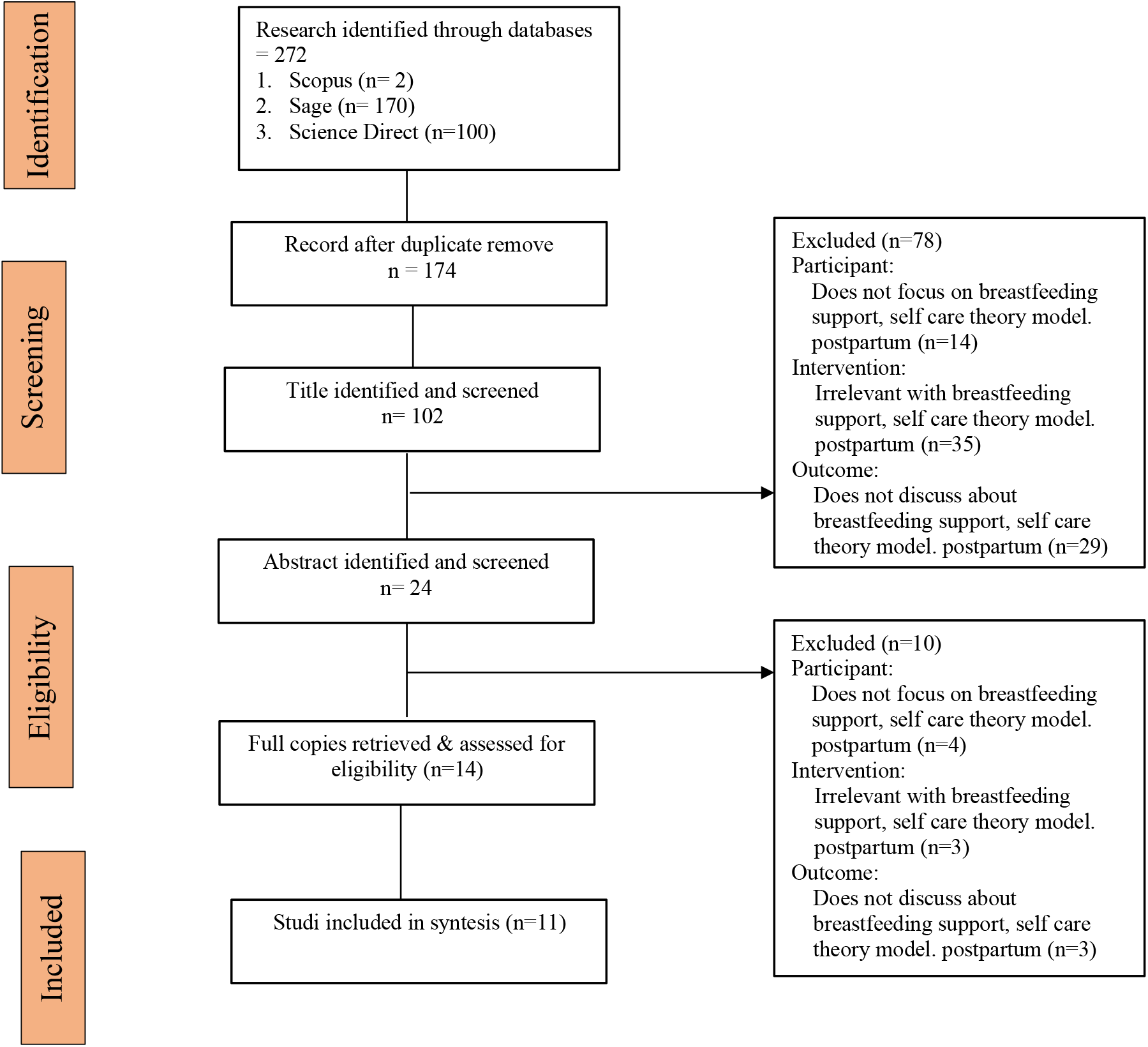
Literature Search Flow Diagram based on PRISMA.

**Table 1.** Results of Article Analysis.

| No | Title, Author, and Year | Metode | Results |
| --- | --- | --- | --- |
| 1 | Aliaa Abd El –Aty<br>Farag Taalab, et all<br>Dependent Care:<br>Applying Orem<br>Self-Care Theory<br>(Taalab et al., 2021)<br>Menoufia Nursing<br>Journal MNJ, Vol. 6,<br>No. 2, Nov 2021,<br>PP:155 -170 | **Design** : A quasi-experimental research design was utilized.<br>**Sample**: A convenience sample of 100 postpartum women.<br>**Variable**: sleep quality and dependent care<br>**Instrument**: An interviewing questionnaire and Orem`s Self-Care Guidelines Checklist were used.<br>**Analysis** chi-square test | The dependent care given by Orem self-care prevent problem related to primiparous postpartum. Conclusion: the care provided using Orem`s self-care model during the postpartum period recovered or prevented the postpartum problems. |
| 2 | Amany Abd El-Fatah Abd<br>El-Azeem Omran, et all<br>Self-Care of Women<br>during Postpartum period<br>in Rural Area (Abd El-Azeem Omran et al.,<br>2020)<br>Egyptian Journal of Health<br>Care, 2020<br>EJHC Vol.11 No.1 | **Design**: Quasi-experimental design<br>**Sample** : 67 postpartum women<br>**Variables**: socio-demographic characteristics, obstetric history Postpartum woman's cultures and beliefs about postpartum period, Postpartum woman's knowledge regarding postpartum self-care, Postpartum woman's self-care during postpartum period, infant's growth and development measurements sheet, home environmental assessment<br>**Instrument**: For data collection three tools were used, 1st tool: an interviewing questionnaire for the postpartum women includes five parts; Part one: socio-demographic characteristics, Part two: obstetric history, Part three: Postpartum woman's cultures and beliefs about postpartum period, Part four: Postpartum woman's knowledge regarding postpartum self-care, Part five: Postpartum woman's self-care during postpartum period, 2nd tool: review of infant's growth and development measurements sheet, 3rd tool: home environmental assessment observational checklist. | The current study revealed that less than three quarter of postpartum women had negative total cultures and beliefs regarding postpartum self-care, more than two fifth of postpartum women had satisfactory knowledge toward total postpartum self-care, also majority had inadequate self-care. With a highly statistically significant difference between knowledge and self-care practice Conclusion: the total score level for self-care practice revealed that the majority of study sample unaware of adequate self-care practice in rural areas. |
|  |  | **Analysis :** Collected data were coded and tabulated using personal computer. Statistical package for social science (SPSS) version 20 was used. Descriptive as well as inferential statistics were used to answer research question. |  |
| 3 | Shimaa Ali Sayed Abdrabo, et all<br>Improvement of Self-care of Postpartum Minor Discomfort (sayed et al., 2022)<br>Assiut Scientific Nursing Journal Vol , (01 ) No, (32), September, 2022, pp (126 - 137 ) Print Issn: 2314-8845 Online Issn: 2682-3799 | **Design:** quasi experimental research design<br>**Sample :** 200 immediate postnatal mother<br>**Variables:** knowledge levels regarding postpartum period and its minor discomforts - Apply health education about postpartum discomfort - Improve women's self-care.<br>**Instrument:** An interviewing questionnaire was developed based on the review of relevant literature to collect maternal personal data and to investigate mother's self-care practices for relieving minor discomfort during postpartum period. It includes: Part I: Addressed personal data such as age, occupation, level of education. Part II: It includes maternal obstetric history as No. of parity, gravidity and current pregnancy data as week of gestation, mode of delivery. Part III: This part assess maternal knowledge<br>**Analysis :** chi- square (cross tabulation) between pre, and post-test after one month, two months and after three months, correlation and one way a nova test were done using computer program (SPSS) | There were a statistical significant difference between knowledge level, self-care pre and post-test after implementing nursing education (P=0.001). Conclusion: there was significant improvement in women's knowledge and reported self-care practice regarding postpartum minor discomfort after health education. Recommendation: Educate mothers in late pregnancy about physiological changes and postpartum minor discomfort and how to deal with this discomfort |
| 4 | Xueli Lei, et all<br>Effects of the PRECEDE-PROCEDE Model on Self-Care Ability and Quality of Life Among Primipara During Puerperium (Lei & Zhou, 2024)<br>Perinatal Nursing and Midwifery– Volume 10: 1–sagepub.com/journals-permissions DOI: | **Design:** Quasi-clinical study<br>**Sample:** 80 case<br>**Variables:** Self-Care and Quality of Life<br>**Instrument:** online questionnaires and scales were used to collect Socio-demographic characteristics, maternal self care ability and quality of life.<br>**Analysis:** Tests such as t-test, Chi-squared test, rank-sum test, and repeated measure analysis of variance | The control group's self-care ability scores were (150.8±9.9), (150.9±9.3), and (152.0±10.2) before intervention, at 3 weeks postpartum, and at 6 weeks postpartum, respectively. For the intervention group, the corresponding scores were (151.1±15.1), (157.8±8.5), and (162.4±7.2). Quality of |
|  | 10.1177/23779608241282392 |  | life scores for the control group were (54.7±8.6), (54.8±7.7), and (55.1±7.7) before intervention, at 3 weeks postpartum, and at 6 weeks postpartum, respectively. At the same time points, while the intervention group saw increases from (55.6±7.6) to (59.2±5.9) and (61.0±5.3). There were statistically significant differences in the time effect and inter-group effect of the total score of self-care ability during puerperium, total score of quality of life, and the score of each dimension between the two groups (P |
| 5 | Shadia Saady Mohamed sayed, et all<br>Effect of Care Provided Using “Orem Self-Care Model” on Postpartum Dependent Care among Primiparous Mothers (Saady Mohamed sayed et al., 2024)<br>Egyptian Journal of Health care, June 2024 EJHC Vol.15 No. 2 | **Design:** A quasi-experimental design<br>**Sample:** 200 pregnant women<br>**Variables:**, knowledge questionnaire form and Orem`s Self-Care Guidelines.<br>**Instrument:** Three tools were utilized for data collection, a structured interviewing questionnaire, knowledge questionnaire form and Orem`s Self-Care Guidelines Checklist.<br>**Analysis:** Cronbach's alpha, Descriptive statistics were applied (e.g., mean, standard deviation, frequency and percentages) | At the post-intervention phase, the women in the study group had more adequate knowledge in comparison with those in the control group. Also, a statistically significant difference the women in the study group had more satisfactory self-care practices in comparison with those in the control group. This was evident in all areas about self-care practices (P value < 0.001) |
| 6 | Reina Dhamanik, et all<br>Application of Orem's Self-Care theory and Pamela's Self-Transcendence to clients with breast engorgement. (Dhamanik et al., 2024)<br>Holistic Nursing Care Approach, Vol 4 No 2, | **Design:** This case study focuses on the application of Dorothea Orem's self-care theory and Pamela G. Reed's Self-Transcendence theory.<br>**Sample:** five cases of breast engorgement in patients with different scales. The cases were taken at two teaching type hospitals. The first and second cases were from a type B hospital and the other case was from a type A hospital. Patients were | The use of Orem's self-care theory and Pamela G. Reed's self-transcendence in two cases of breast engorgement made it easier for health workers in the process of assessment to evaluation. about breastfeeding problems so that patients are able |
|  | Agustus 2024 e-ISSN: 2808-2095 | selected based on the results of breast engorgement scale assessment<br>**Variables:** Breast Engorgement. Physical, emotional, cognitive changes<br>**Instrument:** Breast engorgement scale assessment.<br>**Analysis:** qualitative case study | to independently apply the interventions provided because they are easy and efficient if done correctly. to help the process of releasing breast milk with a condition of breast duct obstruction that can reduce the impact of mastitis and overcome anxiety conditions due to psychological changes in postpartum mothers in the form of providing Evidence Based Nursing Practice A.B.E (Arugaan Breast Engorgement) techniques. where this intervention is able to empower mothers to do massage with emphasis on certain meridian points which have the aim of focusing on the process of smooth breast milk production. |
| 7 | Eman Youssif Ali Awad, et all<br>Effect of prenatal Educational program on Knowledge and Self Care Practices Regarding Prevention of Breast Problems among Lactating Primiparous Women (Eman Youssif Ali Awad Faiza Mohamed EL-Said, 2023)<br>Vol. 28. No.1 Februari 2023 | **Design :** A quasi-experimental research design (one group pre-test, post-test) was adopted for the current investigation.<br>**Sample:** Purposive sample comprised of (100) primipara women during third trimester, intended to breastfeed their infants, free from any breast problems and accepting to participate in the study<br>**Variable:**<br>**Instrument:** For this study's data collection, three instruments were used: Tool I: A-self-administrative questionnaire which composed of two parts, part I: socio-demographic characteristics of the studied women, Part II: Reproductive history of the studied women, tool II: Knowledge assessment tool which composed of two parts, part I: knowledge of primipara women about breast feeding characteristics, Part II: primipara women's knowledge about | The relationship between practice and knowledge was statistically significant (p 0.001*), More than one-third (37%) of the women studied had poor knowledge before the intervention, but the majority (85%) had good knowledge after the intervention. There was also a statistically significant positive correlation between the studied women's knowledge and their practices after the prenatal education program. Conclusion: After the implementation of the prenatal education program, a number of |
|  |  | breast feeding problems and tool III: women's self-care practices regarding prevention of breast problems.<br>**Analysis:** Chi-square test, ANOVA test, T Test | self-care knowledge and practices were significantly improved. Recommendation: It is recommended to establish an antenatal education program for all primiparous women to improve their understanding of breast problems and self-care practices. |
| 8 | Sendedeh Mahboobeh, et al<br>The Effect of Prenatal Self-Care Based on Orem's Theory on Preterm Birth Occurrence in Women at Risk for Preterm Birth (Rezaeean et al., 2020)<br>Iranian Journal of Nursing and Midwifery Research. Volume 25 Issue 3 , May-June 2020 | **Design** : : The present clinical trial was conducted on<br>**Sample:** 176 pregnant women at 24–26 weeks at risk for preterm birth in Mashhad, Iran, from December 2015 to October 2016. A multistage sampling method was used in this study. The intervention group (88 pregnant women) received individual self-care education but the control group (88 pregnant women) received only common prenatal care.<br>**Variable:** incidence of preterm birth<br>**Instrument:** The data collection instruments included a questionnaire for demographic and obstetric characteristics, the Holbrook's preterm delivery screening questionnaire, and the Hart Prenatal Care Actions Scale (HPCAS)<br>**Analysis:** Chi-square test | There was a statistically significant difference between intervention and control groups in terms of preterm birth occurrence (6.80% vs 20.50%) ($\chi^2 = 6.90$, $df = 1$, $p = 0.008$). The incidence of preterm birth in the intervention group was approximately three times higher than that in the control group. Conclusions: Given that educational interventions could reduce the incidence of preterm birth, it is suggested that the women at risk for preterm birth are trained for prenatal self-care |
| 9 | Eti Surtiati, et all<br>The Effect of Dorothy Orem's Self-Care on Independence and Anxiety of Pregnant Women in the III Trimester in Facing Labor (Eti Surtiati dkk, 2020)<br>Vol. 15 No 1 Mei 2023 | **Design** : *Quasi Eksperimen pre test control group design.*<br>**Sample:** pregnant women at the Bogor Health Center with the inclusion criteria of III trimester pregnant women and exclusion of pregnant women with complications, obtained a total of 40 respondents consisting of 20 respondents who received the intervention and 20 respondents as controls..<br>**Variable:** Independence and Anxiety<br>**Instrument:**<br>The anxiety variable questionnaire instrument used was the HARS instrument while the independence | Statistical tests using the t test and chi square statistical tests, between the independence variables, the statistical test results obtained a p value = 0.010 ($p$ value = $<0.05$, at $\alpha = 5\%$), This means that there is a difference in mean anxiety scores in the intervention group and the control group, or it can be said that at $\alpha = 5\%$ |
|  |  | variable used a modified instrument from Harianti's research.<br>**Analysis:** Chi Square and t-test | there is a significant effect on the Dorothy Orem Self-care nursing model on the anxiety of III trimester pregnant women in facing labor. |
| 10 | Eman Abd El-hady, et all<br>Self-Care Practices of Primipara Women Regarding Breast Engorgement (Abd El-hady et al., 2021)<br>Tanta Scientific Nursing Journal Vol. 20 No. 1 February, 2021 | **Design :** Descriptive research design was used in this study.<br>**Sample:** 200 women were included in the study<br>**Variable:** Socio-demographic data of the women, Reproductive history, Knowledge of primipara women regarding breast engorgement, Self-care practices of primipara women regarding breast engorgement, LATCH Breastfeeding Charting Scale<br>**Instrument:** Structured interview schedule, Socio-demographic data of the women, Reproductive history, Knowledge of primipara women regarding breast engorgement, Self-care practices of primipara women regarding breast engorgement, LATCH Breastfeeding Charting Scale<br>**Analysis:** Chi-square test($\chi^2$). non-parametric data of independent sample, Z value of Mann whitney testKruskal-Wallis ($\chi^2$ value Pearson's correlation coefficient (r) | The study revealed that almost three quarters (77.5%) of women had poor level of knowledge followed by (17.0% and 5.5% respectively) of them had fair and good level of knowledge. The entire of the studied women with breast engorgement had unsatisfactory practices. It also illustrated that, only 27.0% had well breastfeeding. |
| 11 | Maryam Chamangasht, et all<br>Efficacy of an Early Self-care-Based EducationProgram on the Self-evaluation of Primiparous Postpartum Mothers: A Randomized Controlled Clinical Trial (Chamangasht et al., 2021)<br>Shiraz E-MedJ. 2021; 22(9):e108132. | **Design :** A randomized controlled clinical trial<br>**Sample:** 58 primiparous women<br>**Variable:** Ethnicity, Women's education level, Spouse's education level , Women's occupation, Spouse's occupation, Family income level, Pregnancy status, Independence status , The medical interventions, Receiving support at household tasks, Receiving support for the care of the infant<br>**Instrument:** A three-part questionnaire consisting of general, developmental, and health deviation postpartum evaluation made by researchers. The intervention group received three education sessions throughout the 3-5, 10-15, and 17-22 days after childbirth. | Before the intervention, there was no statistically significant difference in the mean total self-evaluation scores between the intervention (131.269 12.742) and control (137 9.600) groups (P-value=0.073), but six weeks after delivery, a significant difference was observed in the mean self-evaluation scores between the intervention (149.692 7.625) and control (122.923 11.495) |
|  |  | **Analysis:** Descriptive statistics (such as mean, standard deviation, and absolute frequency) and statistical tests (including t-test, paired t-test, and chi-square) or non-parametric statistical tests (e.g., the Wilcoxon and Mann-Whitney test) | groups (P-value < 0.001). |

## Results and Discussion

The results of a review of 11 articles describing related as one of the applications of breastfeeding based on the Orem Self-care model theory in postpartum mothers, namely research (Dhamanik et al., 2024), (Rezaeean et al., 2020) (Eman Youssif Ali Awad Faiza Mohamed EL-Said, 2023), (Eti Surtiati dkk, 2020), (Abd El-hady et al., 2021), (Taalab et al., 2021) (Chamangasht et al., 2021), (Saady Mohamed sayed et al., 2024), (Lei & Zhou, 2024), (sayed et al., 2022), (Abd El-Azeem Omran et al., 2020).

Orem’s self-care theory emerged as the first concept of self-care as it describes the various actions mothers take to maintain their life, health and well-being. Orem states that everyone has a natural level of self-care ability, and support from family, friends and healthcare professionals can help. The second concept is therapeutic self-care demand, which describes the maternal activities required to achieve a state. It is divided into three concepts: universal self-care, developmental needs, and self-care health. The third concept is the agent of care, which indicates the mother’s ability to perform independently, which is influenced by age, developmental conditions, life experiences, socio-cultural orientation, and medical situation. Overall, Orem’s self-care theory will be disrupted if the first and second concepts are out of balance with the mother’s inability to perform necessary activities. As a result, the mother needs support from others to perform the necessary tasks (such as nurses, midwives, and family).(Lambermon et al., 2020).

### 1. Ontological study of self-care-based breastfeeding support for mothers

The ontological study of self-care-based maternal breastfeeding support focuses on understanding the nature and essence of support that aims to enable mothers to take care of themselves while breastfeeding. Self-care, also known as self-care, is a concept that emphasizes a person’s ability to take control of their own physical and mental health. In this way, breastfeeding support is considered as outside help, but it also helps mothers to take care of themselves during the breastfeeding process.(Lambermon et al., 2020).

In the ontological study of self-care-based breastfeeding support, some of the main aspects that can be studied include: The nature of self-care support for breastfeeding mothers. To support mothers in maintaining their own physical, mental and emotional health, self-care support includes help that goes beyond the physical. Examples of this support may include training practical skills for breastfeeding, education on mental health after childbirth, and stress management techniques.

The essence of empowerment in breastfeeding support ontology: in this context, finding components of support that emphasize maternal empowerment. It is considered valuable when self-care support helps mothers feel more confident and able to overcome breastfeeding difficulties without relying on others. This study emphasizes the importance of support, which not only provides direct assistance, but also helps mothers learn to care for themselves. Independence as part of breastfeeding self-care is linked to maternal independence. Support helps mothers become more independent in caring for their babies and themselves. For example, do mothers feel more confident to breastfeed in public, time their breaks, or recognize signs that they need rest and nutrition.

In the relationship between self-care support and maternal emotional well-being, the presence of self-care support affects the emotional well-being of breastfeeding mothers, including how they manage distress, fatigue, and anxiety. Support that pays attention to emotional health can give moms space to rest and feel good (Eti Surtiati dkk, 2020)

The role of social support in breastfeeding mothers’ self-care although the focus is on self-care, social support is still important in helping breastfeeding mothers learn how to do their own self-care. The ontology of this study also recognizes that independence in self-care does not eliminate the support role of others instead, it helps mothers feel more capable of handling their own needs. Help from a partner or family that allows mothers to have time for themselves, for example, would be considered empowering support.

Values that direct self-care in breastfeeding support such as self-confidence, independence, and self-esteem are the basis of self-care-based support. This ontology study investigates how effective breastfeeding support helps mothers care for and value themselves. Such methods can increase the value of support as they help breastfeeding mothers directly and instill skills and confidence in the long term (Eman Youssif Ali Awad Faiza Mohamed EL-Said, 2023)

Mothers’ subjective experiences of self-care will be the main focus of this ontology study, emphasizing how they perceive the self-care-based support provided. Each mother has unique experiences related to the support that enhances their self-care, how mothers make meaning of these experiences and how they feel about the support.

### 2. Epistemological study on breastfeeding support for self-care-based mothers

Knowledge about self-care acquired, developed and understood in the context of breastfeeding is studied through an epistemological approach to self-care-based breastfeeding support for mothers. This epistemological approach demonstrates the sources, processes, and validity of knowledge related to self-care in supporting breastfeeding mothers, especially enabling them to care for themselves while meeting the needs of their child. It also includes how mothers, health workers and the social environment view self-care as a valuable support.(Lambermon et al., 2020)

Some epistemological elements of self-care-based breastfeeding support include sources of knowledge about self-care for breastfeeding mothers, various sources from which mothers obtain knowledge about self-care practices during breastfeeding, such as medical personnel, family, community, health literature, and social media. The epistemological perspective examines how mothers use these sources to determine their trustworthiness and reliability, and the role of each source in shaping self-care knowledge.(Eman Youssif Ali Awad Faiza Mohamed EL-Said, 2023)

The process of knowledge formation about self-care investigates how knowledge about self-care emerges and develops. Epistemological studies understand how breastfeeding mothers construct an understanding of self-care as an important support in managing the physical and emotional challenges of breastfeeding. This knowledge in the context of breastfeeding may be acquired through personal experience, professional instruction, or advice from fellow mothers.

Criteria for validation and reliability of information about self-care in Breastfeeding mothers receive a variety of information about self-care for their own health. how they evaluate the credibility of this information, including how they verify and select the self-care strategies they consider most effective. To understand how breastfeeding mothers make decisions about what is important and valid in their own self-care, this epistemological approach is essential.(Lambermon et al., 2020)

The role of experience in building self-care knowledge of breastfeeding mothers often relies on personal experience to gain knowledge about self-care. This shows how a mother’s subjective experience influences their perception and understanding of self-care-based support. For example, whether they are more open to new self-care practices due to previous experiences of stress or fatigue, or conversely, whether positive experiences make them more open to self-care.

The influence of values and beliefs in self-care practices Personal and cultural values greatly influence breastfeeding mothers’ understanding and beliefs about self-care. In this epistemological study, how mothers’ individual cultural and social values influence their self-care understandings and practices is examined. For example, whether certain cultures support or discourage mothers from caring for themselves, and how this affects mothers’ perceptions of caring for themselves as part of breastfeeding support.

Social construction of self-care support knowledge about self-care is often a social construction formed through interaction with the community or society. The epistemological study of how society and the surrounding environment influence mothers’ knowledge of self-care during breastfeeding. Family and friends may provide moral or ethical support in some communities for self-care support.

The way self-care practices are understood as part of support in epistemology, self-care support is considered as practical knowledge that mothers can use to manage their well-being during breastfeeding. how mothers understand and apply self-care practices supported by those around them and whether they perceive it as effective help.(Lambermon et al., 2020)

### 3. Axiological study on breastfeeding support for self-care-based mothers

Axiologically, it concentrates on the values and norms associated with self-care practices in breastfeeding. In other words, it investigates the meaning, value and importance of self-care for mothers during breastfeeding from an ethical, moral and social perspective.

Some key aspects of the axiological study of self-care-based breastfeeding support are the value of independence and self-esteem in self-care support self-care-based support for breastfeeding mothers strongly emphasizes these values and how these values are articulated in support that enables breastfeeding mothers to manage their own physical and emotional well-being.(Dhamanik et al., 2024)

Social values that support or hinder self-care Every society has norms and values that can support or hinder breastfeeding mothers’ self-care. For example, values in some communities emphasize that mothers should be completely focused on their babies, which can reduce support for self-care. These social values and how the community sees the importance of maintaining maternal health during breastfeeding.

The ethics and responsibilities of providing self-care support, benefiting mothers and adhering to ethical principles in providing support, are aligned with ethics that respect mothers’ autonomy, including helping them make the best decisions for themselves. This means that support is not only directive, but also respects the mother’s right to choose what they need for their own health.

The emotional value of self-care for breastfeeding mothers’ well-being self-care support has deep emotional value for mothers’ well-being, such as reducing stress, increasing self-confidence, and making space for mental health. Identifying these emotional values, including how self-care helps breastfeeding mothers deal with the daily challenges of caring for their babies.(Saady Mohamed sayed et al., 2024) Community perceptions of self-care as a component of breastfeeding support investigated community perceptions of self-care as a component of breastfeeding support. This support may receive less attention because self-care may be considered less important than the needs of the baby in some cultural contexts. Here, sociology will examine whether self-care is considered important and valuable by society or whether it is the mother’s right to take care of their health during breastfeeding.

Value conflicts between social demands and self-care needs often conflict with self-care support. For example, people believe that breastfeeding mothers will give all their care and attention to their babies without considering their personal needs. This axiological study investigates how mothers deal with and balance these value clashes, as well as how they navigate between meeting social expectations and maintaining self-care as mothers.

The empowerment value in self-care support, is a key value in self-care support for breastfeeding mothers because providing self-care-based support encourages mothers to recognize their own needs and gives them the power to meet their needs. how self-care helps breastfeeding mothers become more independent and better able to make their own decisions..(Lambermon et al., 2020)

## Conclusion

Based on a review of 11 articles discussing infant breastfeeding support from the perspective of the self-care model, more efficient strategies to increase milk production can be developed by understanding the self-care model. In addition, the self-care model can be applied in breasfeeding support by providing self-care to improve breastfeeding practices.

## Data Availability

All data produced in the present work are contained in the manuscript

## Notes

### Competing Interest Statement

The authors have declared no competing interest.

### Funding Statement

This study did not receive any funding

### Summary of Updates

revision of the abstract and tables

